# “They don’t just treat the sickness; they treat you like a whole person”: What patients with pancreatic cancer want their healthcare providers to know

**DOI:** 10.1101/2025.10.06.25337462

**Authors:** Arman Bahmani, Jessica Stern, Maud Celestin-Joachim, Nishita Matangi, Lizbeth Rivas, Betty Ferrell, Susanne Montgomery, Nikhil R. Thiruvengadam

## Abstract

**Objective:** Patients with advanced pancreatic cancer and their families encounter multidimensional challenges that extend beyond disease management, encompassing informational gaps, cultural barriers, and unmet psychosocial needs. Few studies have examined the perspectives of patients and their caregivers on what constitutes effective, compassionate care, especially following the peak of the COVID-19 global health emergency, a time of delayed screening and treatments, partly due to an overburdened healthcare system. The purpose of this study was to identify current gaps and factors responsible for improving quality of life among patients diagnosed with pancreatic cancer and their caregivers.

**Methods:** This qualitative study was conducted in 2024 and 2025 using content analysis. Semi-structured interviews of 21 adults, including patients recently diagnosed with pancreatic cancer (N=10) and family caregivers (N=11) across multiple care settings explored their experiences with pancreatic cancer diagnosis and treatment. Interviews were audio-recorded, transcribed and analyzed with MaxQDA software. Constant comparative analyses to identify major themes and subthemes were used and inductively derived codes were created to develop the conceptual model for patients with pancreatic cancer.

**Results:** The analyses yielded four major themes: (1) Patient experience and medical journey were marked by frequent treatment delays; (2) Healthcare navigation and barriers included systemic delays, financial strain, and depersonalized interactions with the healthcare team; (3) End-of-life conversations and palliative care were often misunderstood, and (4) Suggestions for healthcare providers emphasized the need for a more holistic approach respecting patient agency, with timely provision of practical resources. The resulting themes fit well into an adaptation of the ARC framework.

**Conclusions:** Patients with advanced pancreatic cancer desire care that is holistic, culturally responsive, patient centered and delivered with clarity and compassion. Using the suggested conceptual model, providers can improve experiences by aligning care with patient values, facilitating practical and spiritual needs early and integrating culturally relevant supports. Embedding these practices across the care continuum may enhance quality of life, reduce suffering, and minimize unnecessary healthcare costs among patients diagnosed with pancreatic cancer.

---

Patients diagnosed with pancreatic cancer face multifaceted challenges that extend beyond the management of their underlying disease, encompassing psychosocial, cultural, and systemic factors that shape their experiences within the healthcare system. Just the cost of pancreatic cancer can be a major financial burden (on average, more than $60,000 and an additional $175,000 with surgical procedures)[1]. Across diverse populations, patients consistently express a desire for clinicians to treat them as whole persons, integrating their values, preferences, and social circumstances into care planning [2, 3]. However, persistent disparities in access to timely, high-quality care remain a significant concern for patients with serious illnesses such as pancreatic cancer.

Extensive evidence demonstrates that sociodemographic factors, including race, ethnicity, socioeconomic status, insurance coverage, language, and geographic location, are associated with differences in diagnosis, treatment utilization, and outcomes for patients with serious illnesses such as pancreatic cancer [4–10]. For example, patients from racial and ethnic minority groups, those with lower socioeconomic status, and the uninsured are less likely to receive recommended therapies, experience delays in diagnosis and treatment, and have poorer survival outcomes [4–10]. These disparities are driven by a complex interplay of patient, provider and systemic factors, including structural barriers, implicit bias, health literacy, and variable access to specialized care [5, 10, 11, 12].

In addition to these structural challenges, communication about goals of care and advance care planning is often suboptimal. Many clinicians report inadequate training and discomfort with serious illness conversations, which can result in care that is misaligned with patient preferences [2, 3]. Patients expect clinicians to initiate discussions about their values and wishes, yet these conversations frequency occur late in the disease trajectory or not at all [3]. Fragmentation of care, documentation barriers, and lack of standardized processes further impede the delivery of patient centered care [2].

Addressing these disparities and communication gaps is essential to improving outcomes and ensuring that all seriously ill patients receive care that honors their individual needs and preferences. Ongoing research and quality improvement efforts are needed to identify and implement strategies that promote equity and whole-person care across all patient populations [4, 10].

Over the past decades, there has been an emphasis on improving quality of life for cancer patients at all stages by incorporating pharmaceutical and non-pharmacological supportive care [13]. More recently, a special emphasis has been on improving “living with and beyond cancer” (LWBC) [14]. One of the patient-centered frameworks developed with that in mind is the ARC (Adversity, Restoration, Compatibility) framework. This framework posits that patients go through mainly three phases: Adversity pertains to the distress of dealing with the physical, emotional and social distress of dealing with the diagnosis of cancer and the initial treatment(s) and the life-changing impact of it all. Restoration refers to the readjustments that happen in the life of a person with cancer as a result of the diagnosis and treatment, including relying on social support and the healthcare system. Compatibility, in this framework, refers to the reconciliation and development of a new reality and outlook on life. The model assumes that the natural sequence would consist of diagnosis, followed by primary treatment, then either remission or active and advanced disease. The latter scenario would be followed by subsequent treatments and finish with “end of life” care.

In pancreatic patients, the framework is often compressed within a short time period, as most patients only survive a few months. Therefore, our team felt that there is a need to adjust the model as most patients are diagnosed late the process, and do not have the option of remission and subsequent treatments. Therefore, we propose an adjusted framework (see Figure 1 below).

**Figure 1.**
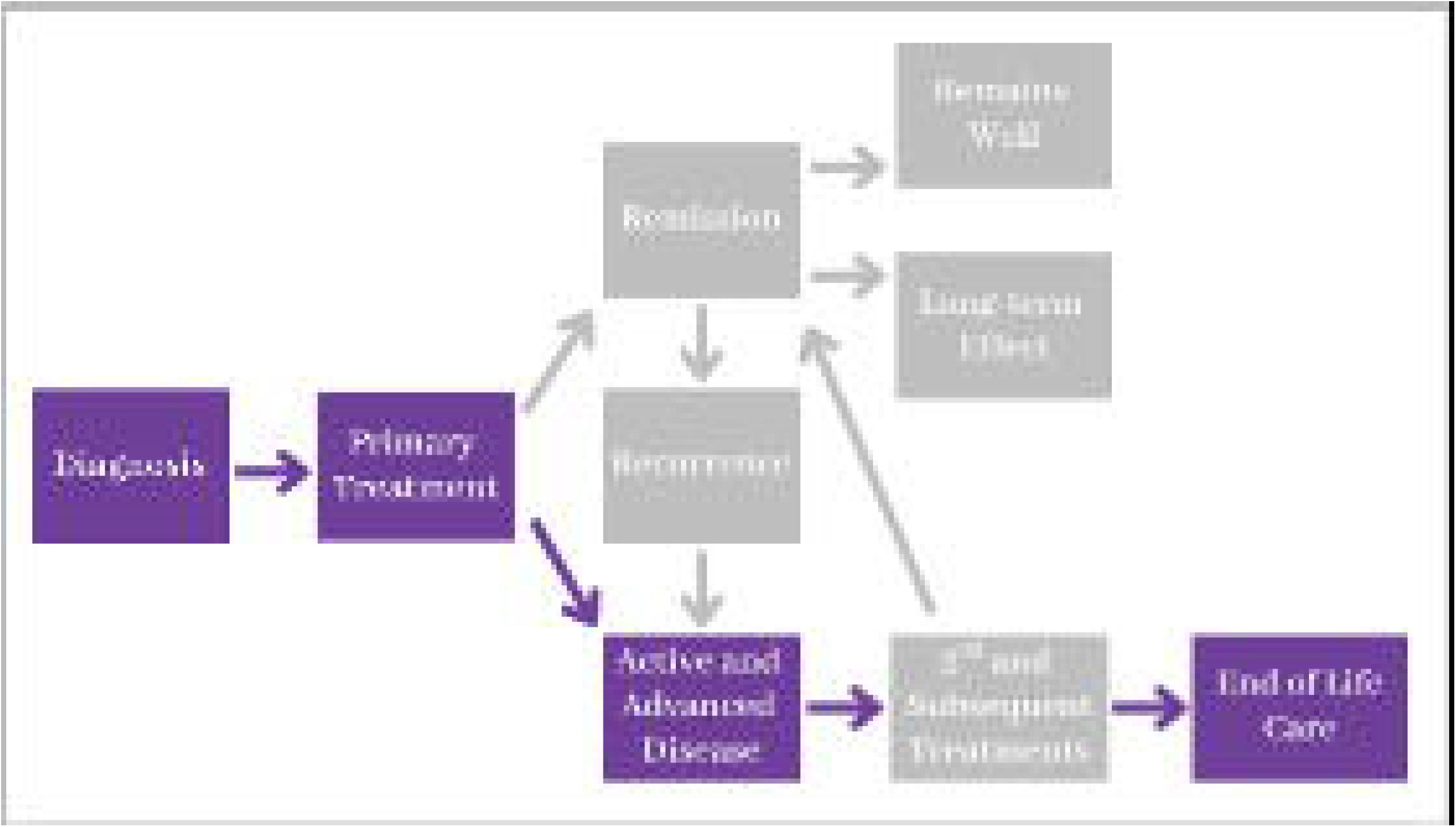
Proposed adapted ARC framework for patients with pancreatic cancer and their caregivers

## METHODS

### Research Design and Sample

This qualitative study was conducted from September 2024 to July 2025 with adult residents of San Bernardino and Riverside Counties in California. Eligibility criteria included being recently diagnosed with pancreatic cancer or serving as a primary family caregiver for someone recently diagnosed. Following diagnosis, participants were invited to participate in a key informant interview to capture early experiences and perspectives.

A total of 21 subjects participated: 10 patients and 11 family caregivers. Of the patient group, 6 were interviewed alone, while the remaining 4 participated in interviews together with one or more family caregivers.

Prior to participation, all subjects provided written informed consent in English or Spanish, which was reviewed and approved by the Loma Linda University Institutional Review Board (IRB# 5240591). Participants received a $10 gift certificate to a local supermarket as a sign of appreciation for their input.

### Data Collection

Semi-structured interviews were digitally recorded and transcribed verbatim by bilingual students and faculty experienced in qualitative analysis. Transcripts were translated from Spanish to English when necessary. Debriefing sessions were held after each interview, and field notes were recorded to capture contextual details. Confidentiality was maintained by de-identifying interviewees and third parties mentioned. The anonymized transcripts were stored permanently at the Loma Linda School of Behavioral Health for research purposes.

### Data Analysis

Transcripts were analyzed using content analysis methodology, with open coding guided by grounded theory principles for emergent themes supported by critical quotes using the 2024 computer software program MaxQDA (version 24.9.1). Two independent coders developed the initial codebook separately, then refined and expanded codes for the final codebook through constant comparative analysis and iterative coding stages. Advanced codes were reviewed collectively to generate an emergent storyline and integrate major themes into a coherent framework adapted from the ARC model. All analyses were conducted in English. Demographic and clinical data information were accessed from the MyChart database and family responses.

### Methodological Considerations

Both individual and joint family caregiver interviews allowed for triangulation of perspective and were included to enhance the depth of analyses. This design facilitated identification of both shared and unique challenges, coping strategies, and unmet needs, consistent with best practices in qualitative research on serious illness and caregiving. The approach also aligns with recommendations to incorporate caregiver voices to provide a more comprehensive understanding of the cancer care journey and its psychosocial impact.

## RESULTS

### Participant Characteristics

Patients (N=10) who were interviewed or whose caregivers were interviewed ranged in age from 51 to 89 years of age, 2/3 were female and 1/3 were males. Five patients conducted the interview in Spanish and the others in English. All caregivers (N=11) except two conducted the interview in English. Ten of the eleven caregivers were related to the patient. The one paid caregiver had worked with the family for 3 years and was considered “family”, therefore, we will refer to caregivers as “family caregivers” in this paper. See Table 1 for details.

**Table 1.** Participants’ demographics.

| MEASURES | Overall group<br>N=21 |  | Patient<br>N=10 |  | Caregiver<br>N=11 |  |
| --- | --- | --- | --- | --- | --- | --- |
|  | n | mean (SD)<br>or % | n | mean (SD) or % | n | mean (SD) or % |
| **DEMOGRAPHIC** |  |  |  |  |  |  |
| **Age** | 21 | 59.29 (13.81) | 10 | 68.20 (11.36) | 11 | 51.18 (10.65) |
| **Gender** | **21** |  | **10** |  | **11** |  |
| Male | 7 | 33.3 | 4 | 40 | 3 | 27.3 |
| Female | 14 | 66.7 | 6 | 60 | 8 | 72.7 |
| **Preferred Language** | 21 |  | 10 |  | 11 |  |
| English | 16 | 76.2 | 8 | 80 | 8 | 72.7 |
| Spanish | 5 | 23.8 | 2 | 20 | 3 | 27.3 |
| **Ethnicity** | **21** |  | **10** |  | **11** |  |
| Non-Hispanic White | 10 | 47.62 | 5 | 50 | 5 | 45.5 |
| Hispanic | 9 | 42.86 | 4 | 40 | 5 | 45.5 |
| Native American | 2 | 9.52 | 1 | 10 | 1 | 9.00 |
| **Interview setting** | **21** |  | **10** |  | **11** |  |
| Alone | 13 |  | 6 | 60 | 7 | 63.63 |
| Joint<br>(accompanied) | 8 |  | 4 | 40 | 4 | 36.36 |

Among the patients four were retired, two were IHSS caregivers (recently disabled), one was unhoused, one was a construction worker. As for the profession of the caregivers, only two were retired (one had been a probation officer). The others were employed: two were nurses, one was a social worker, one was a construction worker, one had a cleaning service, two were homemakers.

The following four themes were identified: 1) Patient experience - medical journey and frequent treatment delays; 2) Healthcare navigation and barriers; 3) End-of-line conversations and care; 4) Recommendations for healthcare providers. Figure 2 below summarizes the themes and subthemes.

**Figure 2.**
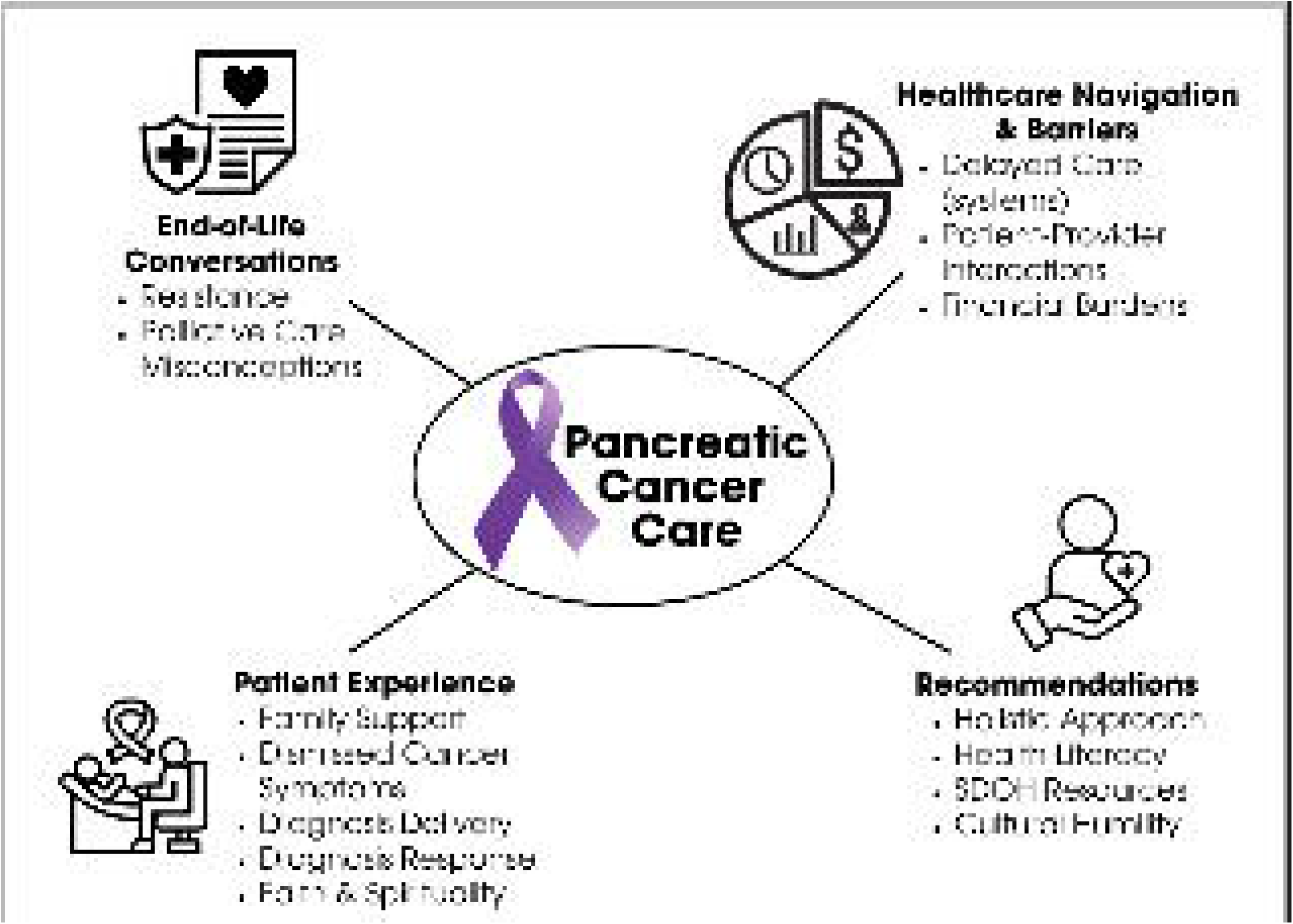
Diagram depicting perspectives of pancreatic cancer patients and their caregivers.

#### Patient experience and medical journey were marked by frequent treatment delays

Prior to diagnosis, pancreatic cancer symptoms such as fatigue and jaundice were often minimized or attributed to less serious causes, leading to delayed attention to symptoms until they became severe enough to warrant an emergency room visit. The circumstances under which participants received the diagnosis varied: while most patients reported receiving these life-altering news in rushed or public settings, compounding psychological distress, a few others were told in private, compassionate environments. Initial reactions to a cancer diagnosis ranged from shock and denial to resignation, acceptance, and determination to “fight,” often shaped by cultural beliefs, and prior experiences with illness. Faith and spirituality were integral to their responses and, for some, the need to appear strong in front of family members to avoid being seen as a burden was a driving force.

Throughout the journey, family was consistently described as the backbone of caregiving, advocacy, and emotional resilience, while strong religious faith, integral to some of our Latino participants, was embraced by patients and caregivers alike as a vital source of coping, strength, and hope during these difficult times. While family support was universally valued, delays in recognizing symptoms frequently hindered timely care initiation. Participants suggested increased education about cancer screenings and symptoms through television, radio and social media as means to lead to quicker diagnosis and treatment initiation.

#### Healthcare navigation and barriers

Language barriers, financial constraints, and limited access to culturally competent care, left the patients and caregivers feeling dismissed, depersonalized, or emotionally unsupported by providers, highlighting the need for compassion, presence, and patient-centered communication. Patients expressed confusion and stress associated with navigating the healthcare system, frequently facing delays in receiving critical care, testing, and treatment due to systemic inefficiencies and poor communication. This contributed to frustration, anxiety, and worsening symptoms. These challenges were compounded by financial hardships, including out-of-pocket costs, insurance denials, and food insecurity, burdens that often became overwhelming for those with limited income or support.

#### End-of-Life conversations and care

There was resistance from the patients to engaging in end-of-life care conversations with healthcare personnel and family caregivers. Cultural beliefs played a significant role in shaping these discussions, influencing comfort levels, timing, and openness. There was also an overarching lack of knowledge about palliative care with some participants confusing palliative care with hospice. Yet, many participants and caregivers expressed a desire for more information and support regarding palliative care options and benefits as they discovered that it could be a potential source of relief, guidance, and improved quality of life during serious illness.

#### Recommendations for healthcare providers

Participants advocated integrating emotional, spiritual, supporting patent and family agency, and practical support alongside clinical treatment. Suggestions for healthcare providers highlighted the importance of a holistic approach in healthcare - looking beyond the disease to care for the whole person’s mind, body, and spirit and “care for the whole person, not just the disease.”

Especially in this fast-progressing illness (late-stage pancreatic cancer) participants wished providers would proactively facilitate timely provision of practical resources for transportation, nutritional support, nursing, and financial assistance early in the care process. They expressed that a lack of simple, clear information often left them feeling lost and fearful. They felt that it would help families feel informed and reassured if verbal explanations were reinforced with clear, jargon-free explanations of disease and treatment pathways, supplemented by visual and accessible educational materials, step-by-step explanations of diagnosis, prognosis, and treatment options, supported by visual aids, printed materials, and other easy-to-understand tools, with “drawings or diagrams” during patient visits. For non-English speakers it was requested that this discussion and related materials available in Spanish or their primary language. This was deemed essential for informed decision-making.

Emotional support and guidance following a cancer diagnosis, before discussing treatment steps, was particularly valued. Patients and caregivers also noted that interactions with medical personnel often felt emotionless and called for increased cultural humility and spiritual education among providers to better support patients’ needs. A culturally attuned approach, featuring language concordant care and sensitivity that respects faith, language, and patient and family agency and roles, was described as foundational to building trust. Spiritual support, including prayer when requested, also emerged as a significant desire among patients who noted that such support could offer hope and faith. Those who expressed a strong desire for this approach and those who experienced such care reported positive interactions with the healthcare system. Results are summarized both in narrative, summary paragraph form as well as in table form with quotes below (see Table 2 below).

**Table 2.**
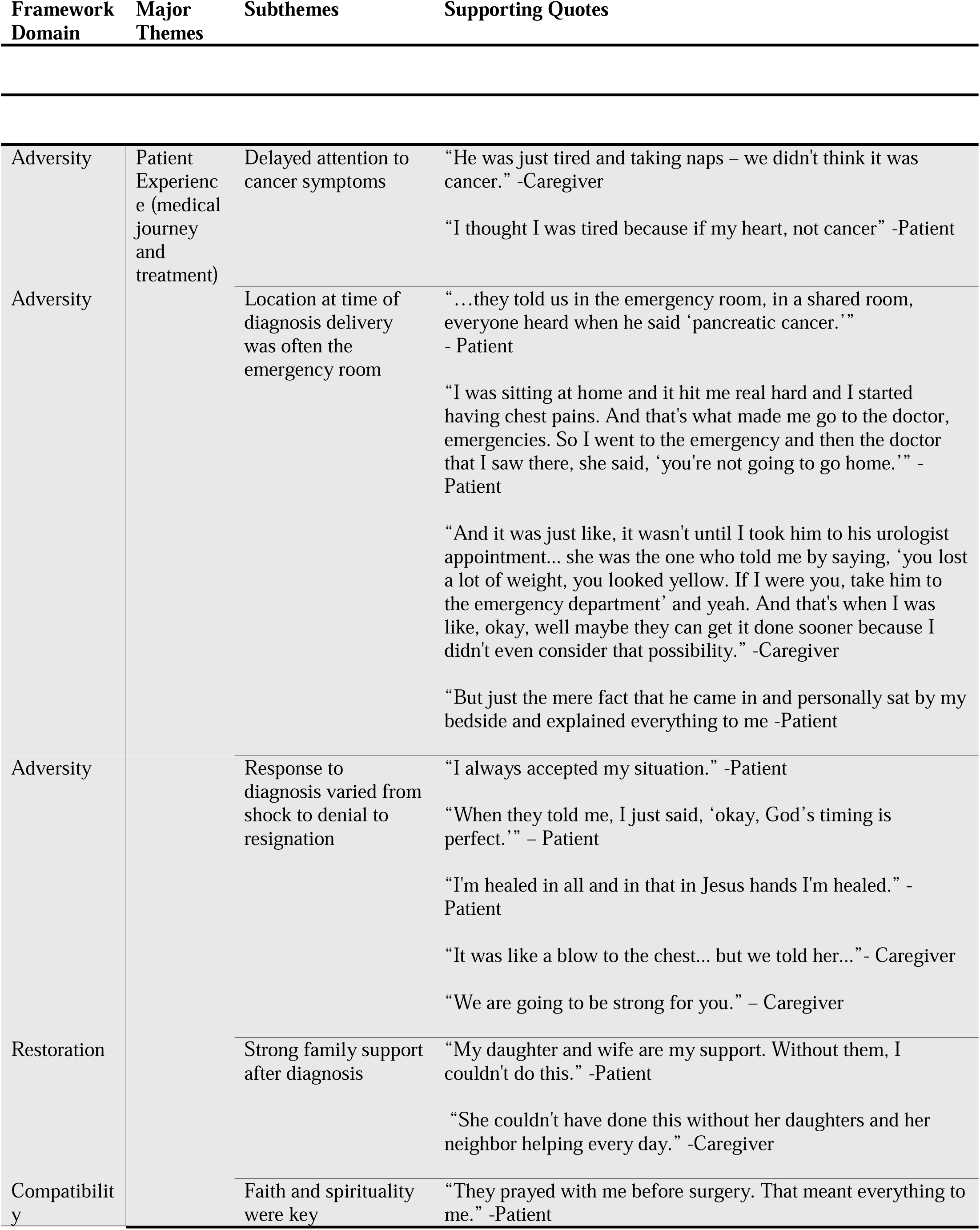
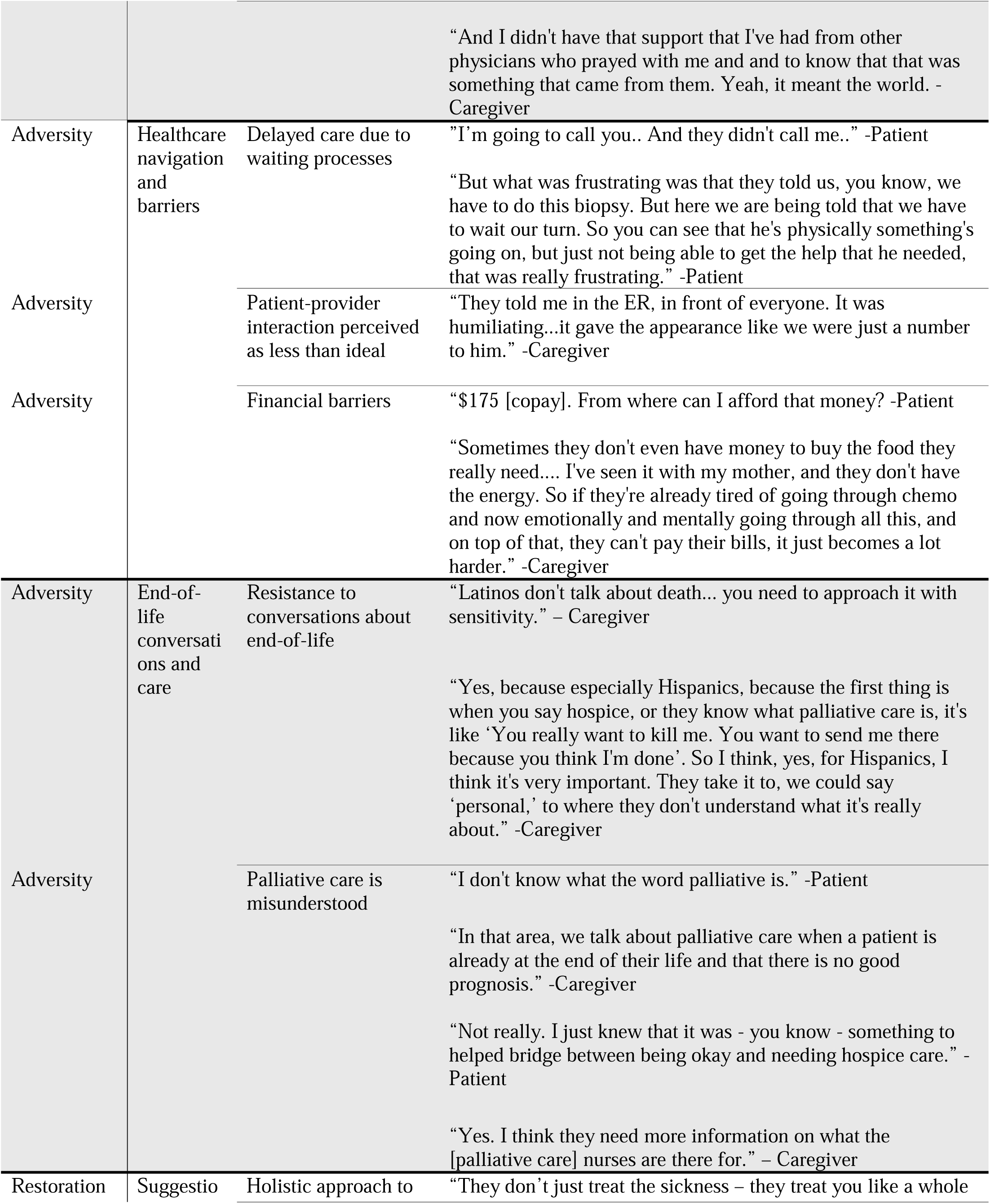
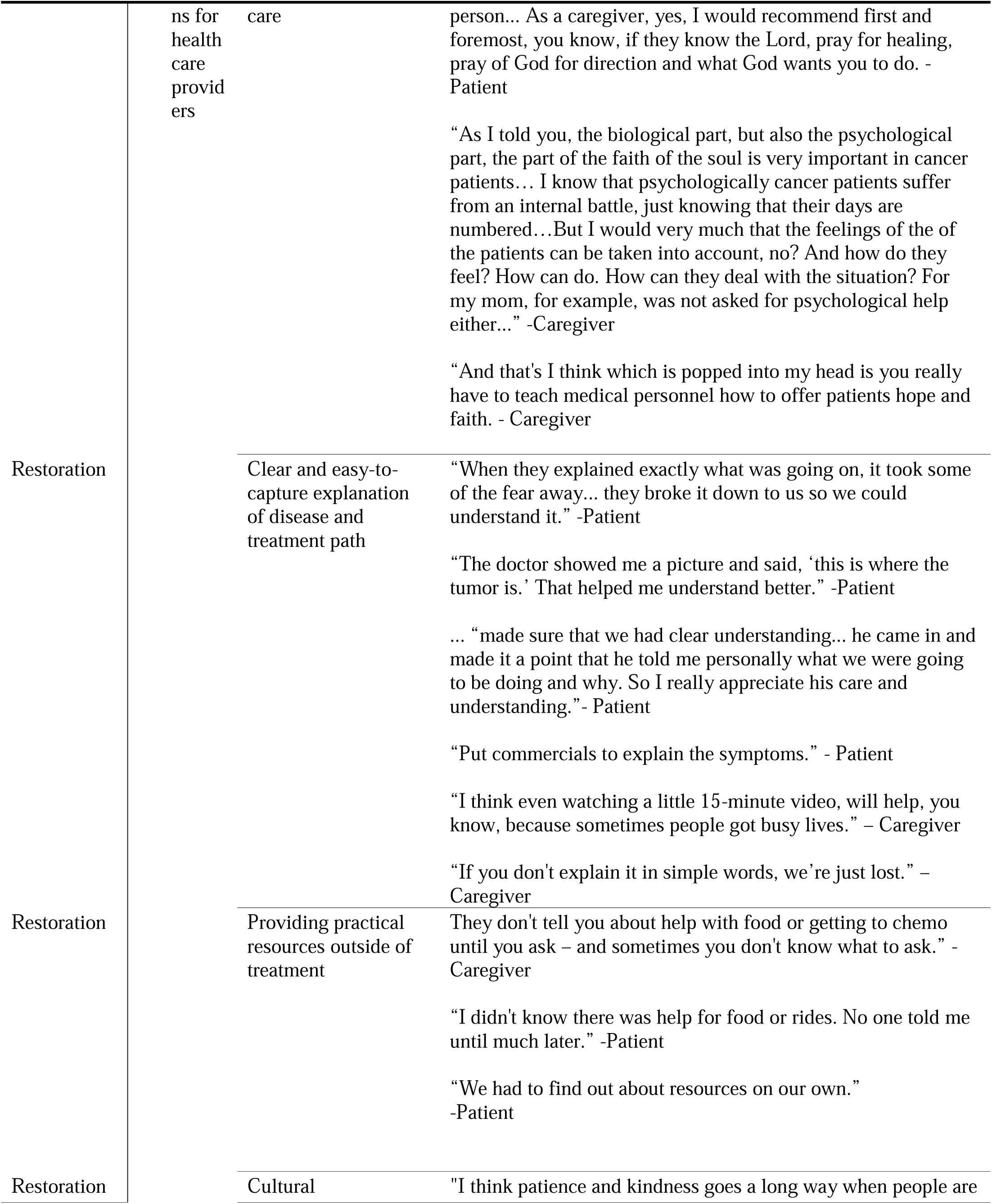
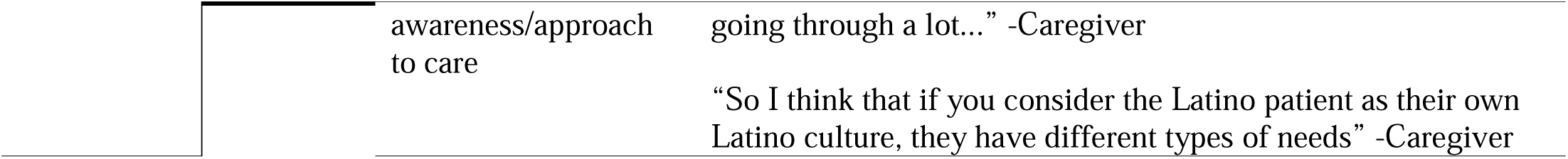
Emergent themes and subthemes from patient and caregiver interviews and corresponding domain concepts.

## DISCUSSION

This study gathered data from our patients and their family caregivers to help identify important factors associated with current health disparities in care and quality of life after a pancreatic cancer diagnosis. Through analysis of patient and family caregiver interviews, four major themes emerged: (1) Medical journey and frequent treatment delays, (2) healthcare navigation and barriers, (3) end-of-life conversations and care, and (4) suggestions for healthcare providers.

The findings emphasize the need for increased access to informative and practical resources, culturally competent care, and holistic support for patients and their caregivers that is respectful of their agency, as they navigate difficult diagnoses and decisions for care.

Delayed recognition and care-seeking for pancreatic cancer symptoms is a well-documented challenge, largely attributable to the nonspecific nature of early symptoms and the absence of effective screening tools. Initial manifestations such as fatigue, mild abdominal discomfort, or weight loss are frequently dismissed by patients as benign or related to aging, stress, or dietary habits, resulting in significant delays before seeking medical attention [15, 16]. Many participants in our study delayed seeking care until more alarming symptoms, such as jaundice, developed, often prompting emergency department visit diagnoses. This pattern is consistently observed in both the literature and patient interviews, with studies highlighting that lack of symptom awareness, perceived provider non-urgency, and patient’s fear or anxiety about a potential cancer diagnosis further contribute to these delays [17, 18].

The majority of participants received their cancer diagnosis in acute or subacute settings, such as the emergency department or urgent care. This finding is consistent with recent studies showing an increasing number of cancer diagnoses in the emergency department [19–22]. These studies also demonstrate the emergency diagnoses are associated with decreased access to primary care, lower socioeconomic status, and worse outcomes, including increased mortality risk [20].

Patients and family caregivers demonstrated a wide range of emotional responses upon receiving a pancreatic cancer diagnosis, including shock, denial, acceptance, and resignation. These reactions reflect the profound psychological impact of confronting a life-threatening illness and the uncertainty it brings. The process of adjustment is highly individualized, influenced by personal beliefs, prior experiences, and the immediate support system. Family caregivers often played a crucial role in helping patients process the diagnosis, fostering resilience and encouraging a sense of collective strength. This variability in emotional response highlights the importance of tailored psychosocial support early in cancer care continuum.

Family support was consistently identified as a cornerstone of coping and adaptation following diagnosis. Patients frequently relied on family members for both emotional reassurance and practical assistance, such as help with daily activities and navigating the healthcare system. Both patients and family caregivers emphasized the importance of these connections for emotional and physical health, which is consistent with evidence linking social support to improved quality of life for cancer patients [23]. These findings reinforce the value of family-centered care approaches in oncology that includes non-stigmatizing referrals to behavioral health sources as part of the care experience.

Faith and spirituality emerged as a significant source of comfort and resilience for many individuals facing pancreatic cancer. Spirituality was also a significant source of strength, whether through personal practice or support from religious leaders. For some, spiritual support from healthcare providers or a referral to non-denominational hospital chaplains was particularly impactful, enhancing their sense of peace and well-being. When patients are receptive, providing spiritual resources and connections can be a positive coping strategy associated with better physical, emotional, and social functioning [24]. The integration of spiritual care into clinical practice, when aligned with patient preferences, can contribute to improved emotional and social outcomes, consistent with literature linking spirituality to better quality of life in serious illness.

Several barriers associated with care and following up after diagnosis were mentioned by patients and caregivers. They frequently experienced delays in care due to long wait times for appointments and complex referral processes which overwhelmed them with many after a while resigning themselves to the process. These barriers often led to increased stress and postponed treatment initiation, particularly for those unfamiliar with the healthcare system. Community health workers and patient navigators have been shown to reduce these delays by assisting with scheduling and care coordination. Such interventions improve timely access to care and patient satisfaction, especially in underserved populations [25, 26].

Communication gaps between patients and providers also emerged as a key barrier to effective care. Many participants described confusion regarding their diagnosis and treatment plans, often exacerbated by the use of medical jargon and a lack of clear, accessible explanations. Patients and caregivers expressed a strong preference for educational materials that are visually engaging, culturally relevant, and available in multiple languages. Evidence supports including short, simple language narrated multimedia education tools, such as *fotonovelas*, narrative films, and bilingual resources, to improve understanding and engagement among underrepresented groups [27]. Enhancing communication through these strategies can empower patients to participate more actively in their care and improve overall satisfaction with the healthcare experience.

Financial barriers extended beyond medical costs to include high copays, cost of living, and for some resulted in their inability to pay for basic necessities for the rest of the family. Many patients complained of out-of-pocket expenses, while caregivers described the added burden of meeting daily needs such as food and utilities during treatment. In some cases, caregivers were the primary breadwinners and faced difficult choices between taking time off work to care for their loved ones or maintaining their income, which further strained family finances and limited their ability to provide support during working days. Patient navigation programs that offer financial counseling and connect families to community resources are essential for reducing these burdens and supporting equitable access to care [25, 26].

Avoidance of end-of-life care discussions was a common theme across all backgrounds, contributing to disparities in access to a utilization of palliative care (PC) and advanced care planning (ACP). Most participants demonstrated limited knowledge of palliative care, often confusing it with hospice, but when this was better explained, expressed interest in learning more about these services, a finding consistent across all racial and ethnic groups [28]. Even among participants who reported understanding palliative care, knowledge gaps persisted. Research shows that African American and Latino patients are less likely to complete advance directives or utilize hospice services, often due to mistrust of the healthcare system, spiritual beliefs, and a preference for family-centered decision-making, while Caucasian patients are more likely to prefer autonomy and comfort-focused care [29–32]. Asian and Hispanic patients also tend to favor family-centered decision-making and have lower rates of advance directive completion, with language and cultural barriers further limiting end-of-life communication and hospice enrollment [33, 34].

Somewhat personalized educational interventions, that also offer culturally adapted materials, including brief videos, and community-based programs, have been shown to significantly improve understanding and acceptance of palliative care and ACP among diverse populations [30, 32–34]. Such targeted educational materials could be integrated into standard care protocols for cancer patients, along with guidance from physicians and community health workers to improve patient understanding and engagement with palliative care services.

Culturally sensitive, physician-led or initiated patient education about ACP has been shown to effectively enhance knowledge and acceptance of these practices among Latino patients with advanced cancer [32]. Culturally adapted educational interventions, including language specific presentations and materials that incorporate effective communication strategies and cultural considerations, have also significantly improved ACP knowledge, as demonstrated by post-intervention assessments [34]. Although our participants recognized the importance of ACP, few had initiated these processes due to the perceived difficulty of engaging in conversations about prognosis and mortality with family members, highlighting the ongoing need for culturally responsive approaches to facilitate these critical discussions. By addressing these needs, healthcare providers can significantly improve the quality of care and outcomes for patients with pancreatic cancer.

Several suggestions given by participants have been reported elsewhere. Although, efforts have been continually implemented throughout the healthcare system to communicate effectively and care for patients, patients and family caregivers in this study consistently emphasized the need for care that is holistic, culturally responsive, and grounded in clear, compassionate communication alongside clinical treatment. Following are evidence-based recommendations that address key domains for improving the quality and equity of care for seriously ill patients with pancreatic cancer (see Table 3.).

**Table 3.**
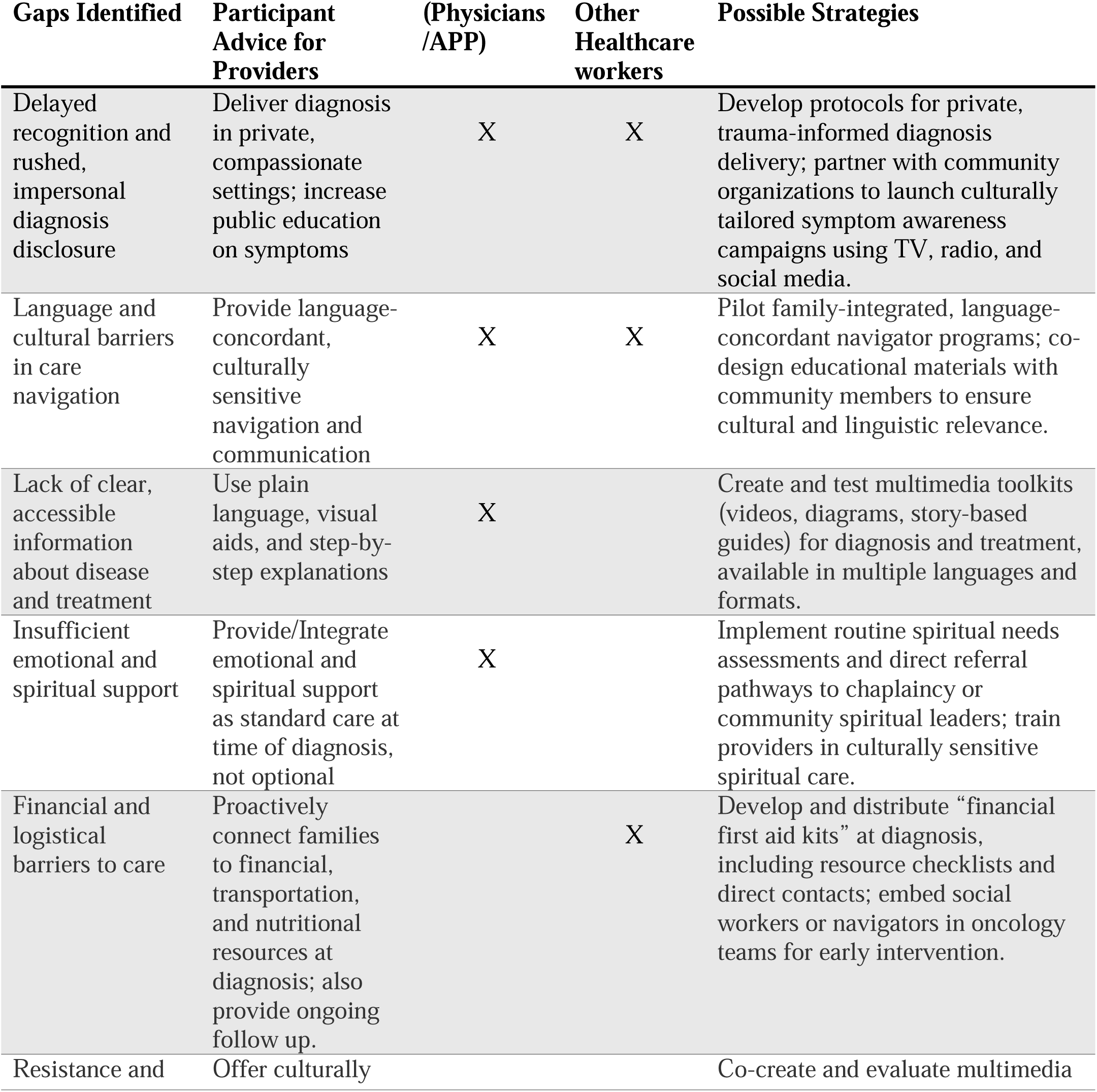
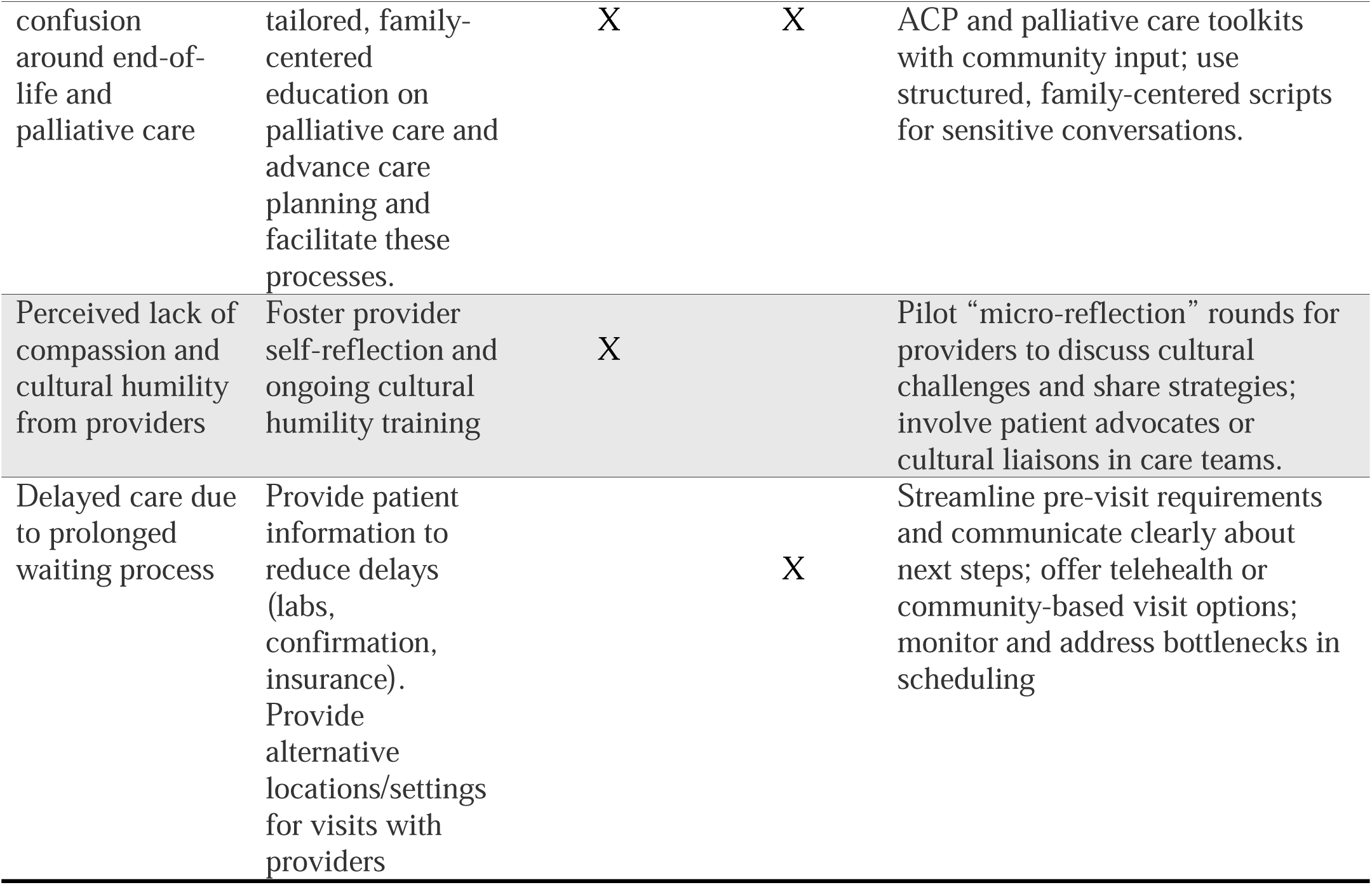
Recommendations and suggested strategies to address identified gaps.

Holistic care requires attention to the physical, psychological, social, and spiritual needs of patients and their families. Those who experienced such care reported positive interactions with the healthcare system, while others expressed a strong desire for this approach. Spiritual support, including prayer when requested, emerged as a significant desire among patients, who noted that such support could offer hope and faith. Prior research supports this shift, showing that culturally tailored, holistic models improve outcomes and patient satisfaction [35–40]. The National Comprehensive Cancer Network (NCCN) specifically highlights the positive impact of spiritual support on quality of life and satisfaction with care, and notes that unmet spiritual needs are common and associated with distress [41]. Other literature recommends early involvement of palliative care specialists for all patients with advanced cancer, regardless of prognosis, to address uncontrolled symptoms, facilitate ACP, and support coping [42, 43].

Multidisciplinary supportive care models that include psychosocial, nutritional, and emotional support have been associated with improved patient and caregiver satisfaction and quality of life; however the subsequent implementation of such promising models have been sparse. Delivery of these approaches remains inconsistent, especially in settings that serve minority populations [44–46]. Further research is needed to develop and implement such interventions for diverse populations.

In our study, participants consistently voiced that they lacked practical, clear information to help them manage their symptoms and self-care. Health literacy is a known determinant of health outcomes and addressing it through better communication strategies remains essential [47].

Limited patient and caregiver health literacy, combined with complex language, leads to worse outcomes, and plain language communication has been recommended as a strategy to improve understanding and engagement [47]. While a plain-language curriculum has been piloted in prostate cancer care, our findings show that this need persists in pancreatic cancer care as of 2025. Information should be delivered incrementally, using plain language and decision aids when appropriate, and providers should ensure adequate time for questions and emotional processing.

Many participants lacked adequate information about the next steps in their care continuum, echoing findings from other qualitative studies that highlight the unmet supportive care needs of pancreatic cancer patients [44]. Providers should offer referrals to financial counseling, transportation assistance, community support organizations, ACD, and disease-specific resources early in the patient’s care.

Another recurring concern was the lack of cultural concordance with providers. Patients reported feeling depersonalized and perceived a lack of compassion, particularly when interacting with providers who did not share or understand their cultural background. Although prior literature has long called for multicultural training among healthcare professionals, our findings suggest that these practices have not been fully adopted [32] and that such training tends to differ by specialty with more such training in primary care. This “disconnect” at the time of a serious and potentially life ending diagnoses can erode trust and complicate discussions around sensitive topics such as ACP. The American Society of Clinical Oncology and other expert groups recommend that clinicians proactively elicit and integrate patients’ cultural values and preferences into care planning assuring their sense of agency at a time when many feel helpless and overwhelmed by the ‘system’, and that health systems invest in workforce diversity and ongoing cultural competence training to address these gaps [48]. These resources and adjustments, if made widely accessible, could enhance symptoms recognition and early care-seeking behavior as well as better outcomes for patients from all backgrounds.

An important extension of the ARC framework in our proposed variation of the original framework is the explicit inclusion of caregivers’ perspectives. Our adapted ARC framework benefits from an added “caregiver layer,” recognizing that holistic cancer care requires attention to family perspectives, especially in preparing for end-of-life transitions. “Adversity” for both patients and caregivers entails dealing with the shock of diagnosis, distress when communication is poor (e.g., hearing news in the emergency room, lack of sensitivity), and the burden of navigating fragmented healthcare systems. Caregivers also highlight financial strain, lack of resources, and stigma around end-of-life conversations. Caregivers not only share in the adversities of delayed diagnoses, poor communication, and financial strain, but also carry a unique burden of navigating healthcare systems, advocating for their loved ones, and managing anticipatory grief. Their voices highlight that “living with and beyond cancer” is not solely a patient experience but a dyadic one, where caregivers’ struggles and strengths profoundly shape the illness trajectory.

Caregivers also play a central role in restoration and compatibility. They mobilize family and community support, seek out resources, and provide the faith and cultural grounding that help patients adapt. At the same time, they reveal gaps in end-of-life communication, describing cultural resistance and misconceptions around palliative care. These insights suggest that they act as the backbone of adaptation, mobilizing support networks, advocating for earlier care, and seeking resources (e.g., food, transport, financial aid). This also emphasizes the need for clear, plain explanations so that, they too, can guide patients through decision-making.

Both patients and caregivers finding meaning through faith, family unity, and cultural values fit with the concept of “compatibility” as described by the ARC framework. They often reframe their role as one of strength, solidarity, and purpose, even as they anticipate grief. Compatibility for caregivers is not only about reconciling cancer as part of life but also reconciling their role as both *supporter* and *individual facing loss*.

From the caregiver perspective, stigma characterized discussions about end-of-life conversations and death. While such conversations occasionally happened, most times the topic was either avoided or dismissed when introduced. Many caregivers do acknowledge the importance of having these discussions, which would make arrangements after the death of the patient smoother, but the close relationships that the caregivers had with their sick loved ones seemed to increase the difficulty of having these end-of-life conversations. One caregiver said that even when her aunt tried having an end-of-life conversation with her, she told her to stop because she did not want to think about that. The importance of having family involved in these end-of-life conversations is crucial but the hesitancy to address these topics could be mitigated by an objective perspective. A study in Hungary found that the general population believed there should be equal engagement of the general practitioner and family member in the end-of-life conversation. Integrating advanced care planning as a standard part of the medical conversation, from the healthcare team, could alleviate both the stigma and the associated emotional complexities. However, when there is resistance, as we found with several of our participants, community health workers – healthcare team members who have earned their trust and have developed a closer relationship with patients – are crucial in moving the process forward before it is too late.

Themes identified in this study were mostly about the construct of “adversity” and then “restoration”. However, only one of the themes fit in the “compatibility” category. This may be due to the short timespan between the diagnosis and the interviews. It could be that participants had not yet reached the stage of “compatibility” where they had developed a new outlook about living with cancer. Or it could be that they were too overwhelmed to advance to that stage. This is one of the challenges that future research among pancreatic patients and their caregivers may need to explore more in depth.

## Strengths and Limitations

Several patients had more than one caregiver. The inclusion of both individual and joint family caregiver interviews allowed for triangulation of perspective and enhanced the depth of thematic analysis. This study has several limitations. The sample size was small and due to the regionality of the hospital, predominantly comprised participants of Mexican descent, all recruited from a single institution. These factors may limit the generalizability of the findings, as variations in sociodemographic backgrounds and healthcare environments could influence experiences and outcomes in pancreatic cancer care. The single-site design may also not fully capture the diversity of barriers and facilitators present in other geographic regions or healthcare systems. Nevertheless, a key strength of this study is the inclusion of caregiver perspectives, which enriches the analysis and offers a more comprehensive view of the complex challenges faced across the cancer care continuum and both our native America and White respondents shared most of the experiences of the noted themes.

## Conclusion

Patients diagnosed with pancreatic cancer face several barriers that are associated with a reduced quality of life. The medical literature demonstrates that targeting training for physicians in serious illness communication and culturally competent care, as well as the integration of multidisciplinary staff such as patient navigators or community health workers, and social workers, can help address gaps in healthcare navigation and support. Implementing culturally sensitive, patient-centered care models and ensuring holistic management can significantly improve the quality of care and quality of life outcomes for patients with pancreatic cancer. System-level changes, including policy reforms and quality improvement initiatives, are essential to promote equitable access and improve outcomes for all patients with pancreatic cancer.

## Data Availability

No data are available in the present study to ensure confidentiality of the interview participants.

